# Evaluation of the second-generation whole-heart motion correction algorithm (SSF2) used to demonstrate the aortic annulus on cardiac CT

**DOI:** 10.1101/2022.06.28.22276816

**Authors:** Yoriaki Matsumoto, Chikako Fujioka, Kazushi Yokomachi, Nobuo Kitera, Eiji Nishimaru, Masao Kiguchi, Toru Higaki, Ikuo Kawashita, Fuminari Tatsugami, Yuko Nakamura, Kazuo Awai

## Abstract

**Purpose:** To investigate the usefulness of the second-generation whole-heart motion correction algorithm (SnapShot Freeze 2.0, SSF2) for demonstrating the aortic annulus at pre-transcatheter aortic valve implantation cardiac CT.

**Method:** We retrospectively analyzed 90 patients with severe aortic stenosis who had undergone cardiac CT on a 256-row CT scanner. The patients were divided into the 3 groups based on their heart rate during the scan (low, < 60 bpm, n = 30; intermediate, 60-69 bpm, n = 30; high, >70 bpm, n = 30). Image datasets were obtained at 40% and 75% of the R-R interval using standard and SSF2 reconstruction. The edge rise distance (ERD) on the CT attenuation profile of the aortic annulus was compared on images subjected to standard- and SSF2 reconstructions. The standard deviations (SD) of area and perimeter were compared using the *F*-test. The image quality was assessed by two observers using a 5-point Likert score.

**Results:** In patients with intermediate and high heart rates, the ERD was significantly shorter on SSF2- than standard reconstructed images (p < 0.01). The SD of area and perimeter were significantly smaller in SSF2 reconstruction than in standard (all: p < 0.05). Except for R-R interval 75% in patients with low heart rate (p = 0.54), the image quality scores were significantly higher for images reconstructed with SSF2 than standard (p < 0.01).

**Conclusions:** For the demonstration of the aortic annulus in patients with high heart rate or a 40% R-R interval, SSF2- was superior to standard reconstruction.

## Introduction

Electrocardiogram-gated cardiac computed tomography (CT) scans are important for planning the transcatheter aortic valve implantation (TAVI) procedure in patients with severe aortic stenosis [1, 2]. However, motion artifacts present a technical challenge because they can compromise the assessment of structures such as the coronary arteries and valves, especially in patients with a high heart rate [3-7]. Inaccurate sizing increases the risk of complications such as perivalvular leak or rupture in TAVI patients [2, 8, 9]. Precise pre-procedural imaging is therefore crucial to assure optimal patient outcome [2, 9]. To avoid motion artifacts, the society of cardiovascular CT guidelines [10] recommend that the heart rate be controlled to be less than 60 beats per minute (bpm) by the oral or intravenous administration of a β-blocker. To correct motion artifacts, technical advances in CT systems have improved the temporal resolution, increased the gantry rotation speed, and applied dual-source CT and multi-segment reconstruction; software solutions have been developed [11].

The first-generation motion correction algorithm (SnapShot Freeze, SSF1; GE Healthcare) is vendor-specific and designed to address coronary motion artifacts on cardiac scans. Its application significantly improved the image quality of the coronary arteries in patients with a high heart rate [12-19]. A second-generation vendor-specific motion correction algorithm (SnapShot Freeze 2.0, SSF2; GE Healthcare) extended the motion correction range to the whole heart within one scan volume [20, 21]. We examined whether the SSF2 algorithm improves the image quality of cardiac CT scans acquired to evaluate not only the coronary arteries but also the aortic valves. We enrolled patients with severe aortic stenosis who had undergone pre-TAVI standard cardiac CT studies without motion correction and pre-TAVI scans subjected to SSF2 reconstruction and compared their image quality.

## Materials and methods

This retrospective study (No. E-2623, Clinical study of motion correction algorithm for cardiac CT) was approved by our institutional review board; informed patient consent for the analyses was waived.

### Study population

We enrolled 108 patients with severe aortic stenosis who underwent cardiac CT as candidates for TAVI between April 2021 and February 2022. Inclusion criteria were patients who underwent contrast-enhanced cardiac CT. Our exclusion criteria were severe renal failure (estimated glomerular filtration rate < 30 ml/min/1.73 m^2^, 15 patients), poor breath holding during scanning (1 patient), extravasation during contrast injection (1 patient) or refusal of CT examination (1 patient). Thus, the final study population consisted of 90 patients; they were 33 male and 57 female ranging in age from 70 to 95 years (median age, 84 years).

To perform stratified analysis of the effect of SSF2 on heart rate during scanning, we divided 90 patients into 3 groups to include the same number of patients according to the relationship between heart rate and image quality [10, 12, 17, 18, 20, 22-24]. In group 1 (n = 30) the heart rate was low (< 60 bpm, range 34 -59 bpm), in group 2 (n = 30) it was intermediate (60 - 69 bpm), and in group 3 (n = 30) it was high (70 bpm or higher, range 70 - 119 bpm).

### CT scanning

All patients were scanned on a 256-detector row CT scanner (Revolution CT; GE Healthcare, Milwaukee, WI, USA); prospective electrocardiogram-gated axial scans were acquired. The scanning-and reconstruction parameters were tube voltage, 120 kVp; tube current, selected by automatic tube current modulation (Smart-mA, GE Healthcare) based on the scout image; noise index, 25; detector collimation, 256 × 0.625 mm or 224 × 0.625 mm depending on the patient’s heart size; gantry rotation, 0.28 seconds; slice thickness, 0.625 mm; scan field of view, 360 mm; display field of view, 200 mm; matrix, 512 × 512; reconstruction, half; reconstruction kernel, HD standard; reconstruction method, deep learning image reconstruction (TrueFidelity, strength High; GE Healthcare) [25-28]. The padding range was 0 - 100% of the R-R interval when a heart rate of less than 60 bpm was recorded during pre-examination monitoring; when it exceeded 60 bpm or was variable. In the presence of arrhythmia the padding range was 0 - 250%. All scans were craniocaudal from the tracheal bifurcation to the level of the inferior margin of the cardiac apex. All patients were able to perform breath-holds during the examination. To achieve high image quality with minimal radiation doses, patients with a heart rate above 60 bpm 5 minutes before the start of scanning were given 2 - 10 mg propranolol hydrochloride (Inderal; Taiyo Holdings Co., Ltd., Tokyo, Japan).

The contrast medium (iodine concentration 350 mg/ml; Iomeron-350; Eisai Co., Ltd., Tokyo, Japan) was injected in triple-phase through a 20-or 22-gauge catheter into the antecubital vein using a power injector (Dual Shot type GX; Nemoto Kyorindo, Tokyo, Japan). The iodine dose of 273 mg/kg in the first phase was administered in 13 seconds. This injection was followed at a speed of 5 seconds by a 50/50 mix of contrast medium (53 mgI/kg) and saline, and finally 100% saline was delivered at the same injection speed. The scanning delay was determined with a bolus-tracking method. A round, approximately 400 mm^2^ region of interest (ROI) was placed in the center of the left atrium and left ventricle, respectively. Scanning was started manually 1 second after contrast enhancement exceeded a predefined threshold of 300 Hounsfield units.

### Data processing

Similar to the SSF1 algorithm [17, 19], the SSF2 algorithm uses data from adjacent cardiac phases (64 milliseconds before and after the target phase) to characterize and correct the motion. The SSF2 algorithm, a fully automated technique based on information and feedback obtained from SSF1 scans, seeks each region at all image volumes for a local path that is consistent with the subset of measured data. Once the vessel’s motion path is identified, the data are discretized into a series of datasets based on when the corresponding projection rays were measured. Each volume dataset in the series undergoes spatial deformation by the motion field. This allows the motion state to be mapped from the respective time to the central reference time, which is determined by the prescribed cardiac phase [29].

All images were reconstructed using the standard (without motion correction) algorithm with deep-learning image reconstruction for reducing the image noise [25-28]. For the cardiac phase, the systolic-(R-R interval, 40%) and diastolic phase (R-R interval, 75%) used for pre-TAVI cardiac CT measurements were selected [2, 4, 29, 30]. As the systolic-and diastolic phases were additionally subjected to SSF2 reconstruction, 4 datasets were obtained for each patient. They were anonymized and transferred to the workstation (Advantage Workstation 4.7, GE Healthcare) for later analysis.

### Quantitative evaluation

The attenuation effect elicited by motion artifacts was analyzed at the aortic annulus. All images were inspected by one radiological technologist (Y.M. with 15 years of experience with cardiac CT studies). To assess the aortic annulus, only axial-and 2D double-oblique multiplanar reconstruction (MPR) images were examined. The aortic annulus was defined as a virtual ring formed by joining the basal attachments of the aortic leaflets [2, 31].

#### Edge rise distance

We generated a 3-directional CT attenuation profile (anterior-, superior-, and inferior direction) of the aortic annulus (Fig 1) using the particle analysis tool (Plot Profile) on the workstation (Ziostation2, Ziosoft, Tokyo, Japan). Areas of calcification where CT attenuation fluctuates significantly were carefully avoided. The CT attenuation profiles were generated at precisely the same location for images reconstructed with standard and SSF2. We cut off the bottom and top 10% of the profile and measured the 10 - 90% edge rise distance (ERD) [32, 33]. The ERD was examined in three directions of the aortic annulus and the mean values were compared on standard-and SFF2 images.

**Fig 1.**
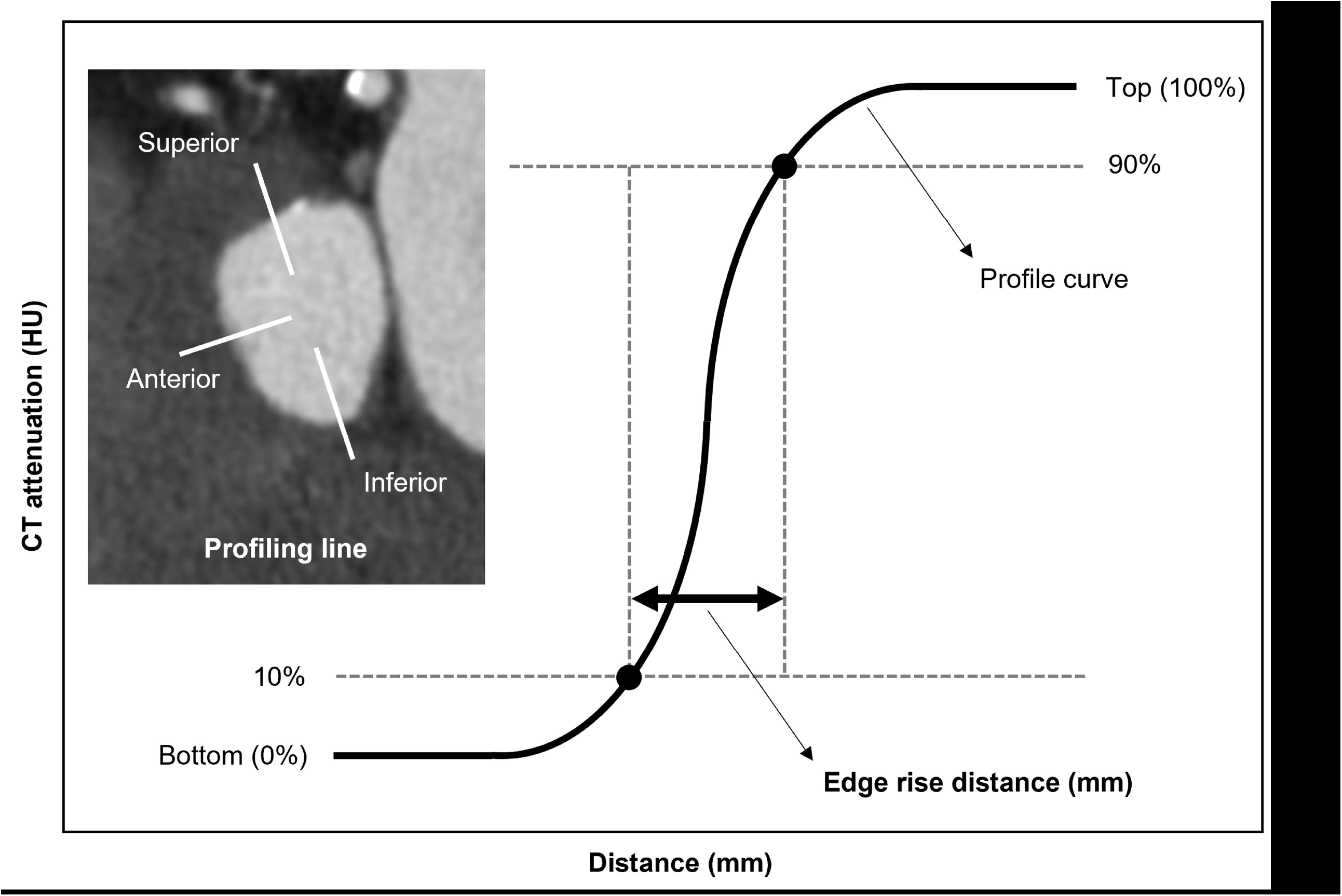
Sample image of ERD. Profile curve of the aortic annulus. The ERD at a pixel attenuation from 10% to 90% of the maximum CT attenuation is shown. CT = computed tomography; HU = Hounsfield units; ERD = edge rise distance

#### Dispersion of sizing

With respect to the sizing of the aortic annulus, we evaluated the dispersion between the two reconstructions. All images were analyzed by two radiological technologists (Y.M. and C.F., with 15 and 18 years of experience in cardiac CT imaging, respectively). They were blinded to presence of SSF2 technique and manually measured the aortic annulus area and perimeter of all patients independently on a CT workstation (Ziostation2, Ziosoft, Tokyo, Japan).

### Contrast-to-noise ratio

To investigate the potential effect of SSF2 reconstruction on the quantitative ERD measurements, we inspected axial images and recorded the CT number and image noise [standard deviation (SD) of the CT number] in a circular ROI placed in the ascending aorta and septal wall of the ventricle. The size of the circular ROI cursor was as large as allowed by the diameter of the ascending aorta (approximately 5.0 - 10.0 mm^2^) and of the septal wall of the ventricle (approximately 1.5 - 3.0 mm^2^). Based on the obtained values we also calculated the contrast-to-noise ratio (CNR) using the formula: (CT number of the ascending aorta minus the CT number of the septal wall of the ventricle) divided by the image noise of the ascending aorta [34].

### Qualitative analysis

Two radiological technologists (Y.M. and C.F., with 15 and 18 years of experience in cardiac CT imaging, respectively) were blinded to presence of the SSF2 technique. They subjectively and independently inspected the MPR images from the sinotubular junction to the left ventricular outflow tract of the datasets for motion artifacts at the aortic annulus level. To grade the image quality they used the 5-point Likert scale where 1 = very poor (motion artifacts resulting in poor visualization of the aortic valve anatomy, not evaluable), 2 = poor (degraded visualization of the aortic valve anatomy due to motion artifacts, not evaluable), 3 = fair (minor motion artifacts with clear delineation of the aortic valve anatomy), 4 = good (no motion artifacts with confident identification of the aortic root anatomy including the cusp nadirs and annular contours), and 5 = excellent (outstanding image quality with a high level of diagnostic certainty with regard to the aortic valve cusps, the leaflet nadirs, and the detection of the aortic annular contours) [29]. Interobserver disagreement was resolved by consensus.

### Statistical analysis

Continuous variables of demographic data, ERD, CT number, image noise and CNR are expressed as the median and range or as percentages or counts, aortic annulus area and perimeter or image quality scores as the mean and SD. The results of ERD, CT number, image noise, CNR and image quality scores were compared on images reconstructed with standard and SSF2 using the Mann-Whitney *U*-test. To compare the dispersion (SD) of area and perimeter between the two reconstructions we used the *F*-test. To determine whether the CNR was equivalent in standard and SSF2 reconstructions, we performed the equivalence test [35]. As the SD of the CNR between the proximal coronary arteries and the adjacent perivascular tissue was 5 in our earlier study [33], we adopted 5 as the equivalent margin. Interobserver agreement in the qualitative evaluation was classified as evaluable (score 3 - 5) and non-evaluable (score 1, 2) assessed with the Cohen kappa κ coefficient where a κ value of less than 0.20 = poor, 0.21 - 0.40 = fair, 0.41 - 0.60 = moderate, 0.61 - 0.80 = substantial, and 0.81 - 1.00 = near perfect agreement. All statistical analyses were performed with JMP 16 (SAS Institute Inc., Cary, NC, USA). Differences of p < 0.05 were considered statistically significant.

## Results

### Patient demographic data

As shown in Table 1, the median overall heart rate during CT image acquisition was 64 bpm (range: 34 - 119 bpm). Of the 90 atients, 70 were in sinus rhythm and 20 exhibited arrhythmias (atrial fibrillation, 19 patients; premature atrial contraction, 1 patient).

**Table 1.** Patient characteristics.

|  | Overall | Patients with low<br>HR (<60 bpm) | Patients with<br>intermediate HR<br>(60-69 bpm) | Patients with<br>high HR (>70<br>bpm) |
| --- | --- | --- | --- | --- |
| Number of patients | 90 | 30 | 30 | 30 |
| Age (years) | 84 (70-95) | 85 (70-95) | 84 (70-91) | 84 (74-94) |
| Male, n (%) | 33 (37%) | 14 (42%) | 11 (33%) | 8 (25%) |
| Height (cm) | 151 (130-169) | 153 (137-167) | 149 (130-169) | 150 (135-169) |
| Body weight (kg) | 51 (33-76) | 50 (33-68) | 53 (33-73) | 52 (35-76) |
| Body mass index (kg/m<sup>2</sup>) | 22.4 (13.6-36.9) | 21.7 (13.6-26.6) | 22.9 (15.5-36.9) | 22.7 (15.8-34.6) |
| estimated glomerular filtration rate (ml/min/1.73 m<sup>2</sup>) | 51.0 (30.3-115.5) | 46.9 (30.3-115.5) | 50.6 (31.3-87.3) | 52.9 (31.0-82.8) |
| Heart rate during the scan (beats/min) | 64 (34-119) | 53 (34-59) | 64 (60-69) | 80 (70-119) |
| No. of patients with arrhythmias during the scan, n (%) | 20 (22%) | 6 (30%) | 3 (15%) | 11 (55%) |
| Atrial fibrillation, n (%) | 19 (21%) | 6 (31%) | 3 (16%) | 10 (53%) |
| Premature atrial contraction, n (%) | 1 (1%) | 0 (0%) | 0 (0%) | 1 (100%) |
| No. of patients using propranolol hydrochloride, n (%) | 69 (77%) | 17 (25%) | 23 (33%) | 29 (42%) |

Values are the median (range) or the number of patients (%).
Propranolol hydrochloride was administered to reduce the heart rate before imaging.

### Quantitative evaluation

#### Edge rise distance

We analyzed 1080 ERDs (3 directions × 4 datasets × 90 patients). The ERD measurement results are presented in Table 2. In atients with a low heart rate, the ERD obtained with standard and SSF2 reconstruction was not significantly different (R-R 40% and -R 75%: p > 0.05). However, in patients with an intermediate heart rate, the ERD at R-R 40% was significantly shorter on SSF2 (2.0 m)- than standard (2.4 mm) images (p < 0.001). In patients whose heart rate was high, the ERD at R-R 40% and R-R 75% was ignificantly shorter on SSF2-than standard images (p < 0.001).

**Table 2.** Comparison of the edge rise distance (mm) on scans subjected to standard-and SSF2 reconstruction.

|  | R-R<br>interval | Standard | SSF2 | *P* |
| --- | --- | --- | --- | --- |
| Patients with low HR (<60 bpm) | 40% | 1.8 (1.0-4.2) | 1.6 (0.9-4.0) | 0.067 |
|  | 75% | 1.9 (1.0-4.2) | 1.8 (0.9-4.1) | 0.122 |
| Patients with intermediate HR (60-69 bpm) | 40% | 2.4 (0.9-5.1) | 2.0 (0.9-4.5) | <0.001 |
|  | 75% | 2.1 (1.0-5.1) | 2.1 (0.9-4.1) | 0.077 |
| Patients with high HR (>70 bpm) | 40% | 2.5 (1.1-5.7) | 2.0 (1.0-5.2) | <0.001 |
|  | 75% | 2.5 (1.1-5.9) | 2.0 (1.0-4.9) | <0.001 |

HR = heart rate. Values are the median (range).

#### Dispersion of sizing

As shown in Table 3, the SD of the aortic annulus area was significantly smaller in SSF2 reconstruction than in standard at low (94.7 vs 63.3 and 105.2 vs 78.9)-, intermediate (71.8 vs 47.9 and 90.4 vs 58.3)-, and high heart rate (58.7 vs 45.1 and 70.3 vs 45.8) R-R interval of 40% and 75% (all: p < 0.05).

**Table 3.** Comparison of SD of the aortic annulus areas (mm^2^) of scans subjected to standard-and SSF2 reconstruction.

|  | R-R<br>interval | Standard | SSF2 | *P* |
| --- | --- | --- | --- | --- |
| Patients with low HR (<60 bpm) | 40% | 448.7 (94.7) | 436.5 (63.3) | 0.002 |
|  | 75% | 428.9 (105.2) | 435.0 (78.9) | 0.029 |
| Patients with intermediate HR (60-69 bpm) | 40% | 442.1 (71.8) | 435.5 (47.9) | 0.002 |
|  | 75% | 445.8 (90.4) | 439.4 (58.3) | 0.001 |
| Patients with high HR (>70 bpm) | 40% | 437.4 (58.7) | 435.8 (45.1) | 0.002 |
|  | 75% | 432.3 (70.3) | 414.6 (45.8) | 0.001 |

HR = heart rate, bpm = beats per minute. Values are the mean (SD).

As shown in Table 4, the SD of the aortic annulus perimeter was also significantly smaller in SSF2 reconstruction than in standard at low (11.6 vs 7.4 and 9.5 vs 6.0)-, intermediate (9.4 vs 5.6 and 10.8 vs 6.8)-, and high heart rate (8.4 vs 4.3 and 9.3 vs 5.4) R-R interval of 40% and 75% (all: p < 0.001).

**Table 4.** Comparison of SD of the aortic annulus perimeter (mm) of scans subjected to standard-and SSF2 reconstruction.

|  | R-R interval | Standard | SSF2 | *P* |
| --- | --- | --- | --- | --- |
| Patients with low HR (<60 bpm) | 40% | 75.0 (11.6) | 75.6 (7.4) | <0.001 |
|  | 75% | 75.4 (9.5) | 74.7 (6.0) | <0.001 |
| Patients with intermediate HR (60-69 bpm) | 40% | 74.6 (9.4) | 74.0 (5.6) | <0.001 |
|  | 75% | 77.0 (10.8) | 75.7 (6.8) | <0.001 |
| Patients with high HR (>70 bpm) | 40% | 74.8 (8.4) | 73.3 (4.3) | <0.001 |
|  | 75% | 71.5 (9.3) | 71.3 (5.4) | <0.001 |

HR = heart rate, bpm = beats per minute. Values are the mean (SD).

#### Contrast-to-noise ratio

As shown in Table 5, the CT number of the ascending aorta and the septal wall of the ventricle and the image noise of the ascending aorta showed no significant difference between the two reconstructions, irrespective of the patients’ heart rate (all: p > 0.05). In addition, these CNR also showed no significant difference between the two reconstructions at low (18.5 vs 19.5, p = 0.404)-, intermediate (16.5 vs 16.3, p = 0.860)-, and high heart rate (17.6 vs 18.1, p = 0.312). The 95% confidence interval for the difference between standard and SSF2 reconstruction was − 3.0 to 1.2 in patients with a low heart rate, − 2.5 to 2.1 in patients with an intermediate heart rate, and − 2.7 to 0.9 in patients with a high heart rate. Because the 95% confidence interval did not cross the bilateral predefined equivalence margin (Fig 2) in all heart rate classes, we considered CNR to be equivalent among our standard and SSF2 reconstitution irrespective of their heart rate.

**Table 5.** CT number, image noise and contrast-to-noise ratio at each site.

|  |  | Standard | SSF2 | *P* |
| --- | --- | --- | --- | --- |
| Patients with low HR (<60 bpm) | CT number of the ascending aorta (HU) | 401.7 (204.7-478.9) | 400.7 (205.0-478.9) | 0.928 |
|  | Image noise of the ascending aorta | 16.9 (13.1-22.3) | 16.3 (11.6-22.2) | 0.206 |
|  | CT number of septal wall of the ventricle (HU) | 83.4 (59.5-116.0) | 85.6 (56.7-115.0) | 0.601 |
|  | Contrast-to-noise ratio | 18.5 (8.5-24.1) | 19.5 (9.0-26.9) | 0.404 |
| Patients with intermediate HR (60-69 bpm) | CT number of the ascending aorta (HU) | 375.0 (308.8-490.0) | 380.6 (299.0-492.5) | 0.962 |
|  | Image noise of the ascending aorta | 17.3 (13.0-26.5) | 17.7 (13.0-27.5) | 0.818 |
|  | CT number of septal wall of the ventricle (HU) | 81.1 (55.9-111.1) | 82.5 (49.3-115.0) | 0.904 |
|  | Contrast-to-noise ratio | 16.5 (11.5-30.8) | 16.3 (9.9-31.0) | 0.860 |
| Patients with high HR (>70 bpm) | CT number of the ascending aorta (HU) | 400.0 (314.5-531.7) | 400.0 (295.5-528.7) | 0.885 |
|  | Image noise of the ascending aorta | 18.1 (15.5-23.5) | 17.0 (13.9-22.0) | 0.161 |
|  | CT number of septal wall of the ventricle (HU) | 80.5 (59.5-117.4) | 83.4 (56.7-123.2) | 0.982 |
|  | Contrast-to-noise ratio | 17.6 (11.7-24.0) | 18.1 (13.3-24.0) | 0.312 |

HR = heart rate, HU = Hounsfield units. Values are the median (range).

**Fig 2.**
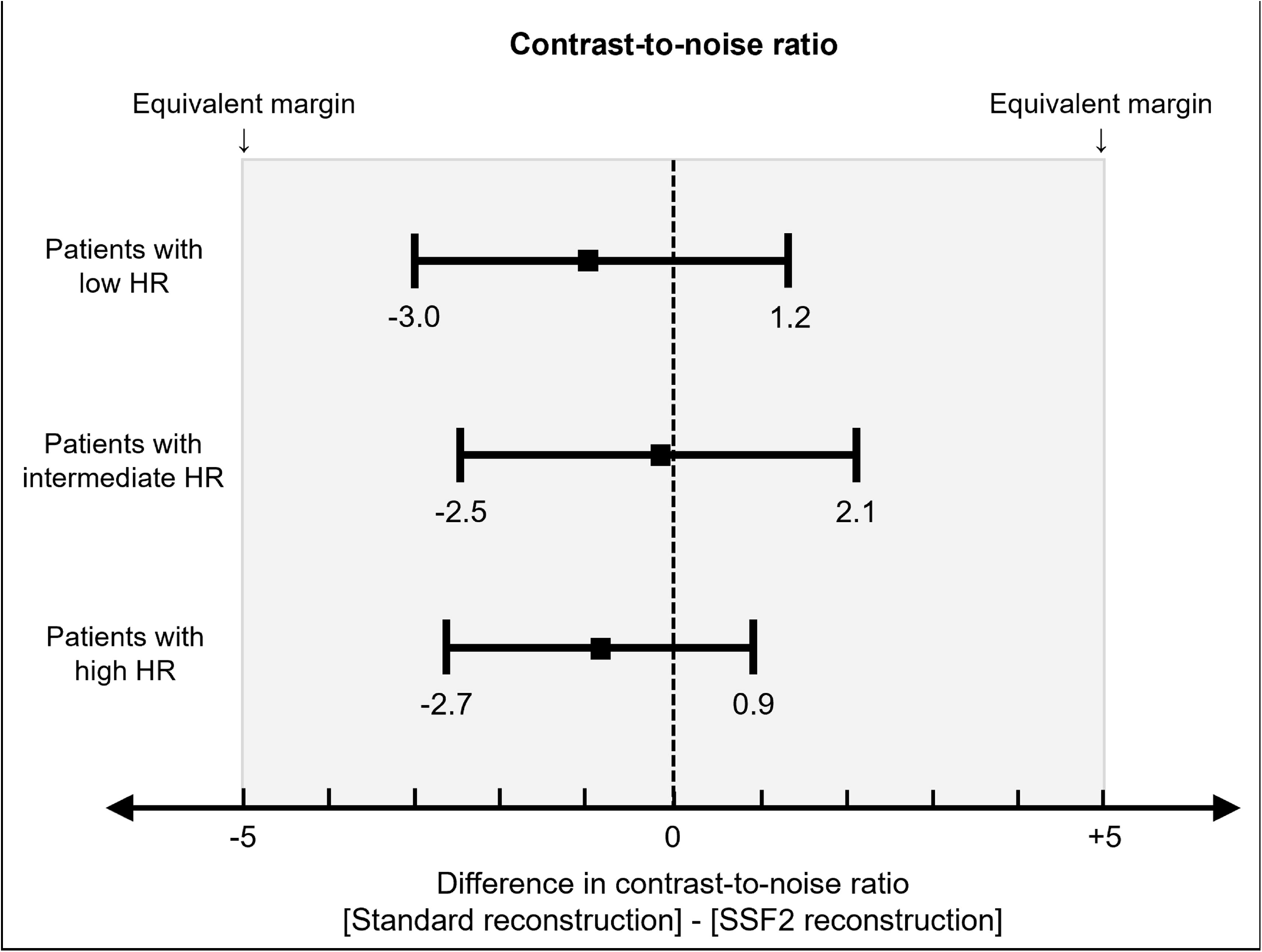
Results of the equivalence test. Results of the equivalence test for the difference in CNR between standard and SSF2 econstruction. CNR = contrast-to-noise ratio; HR = heart rate; SSF2 = SnapShot Freeze 2

### Qualitative analysis

Table 6 shows the results of the visual evaluation of MPR images submitted by our two readers. In patients with a low heart rate, at R-R 75%, there was no significant difference in the mean image scores assigned to images subjected to standard-or SSF2 reconstruction (p = 0.540). At R-R 40% the visualization scores were significantly higher for images reconstructed with SSF2 than standard (all: p < 0.01). There was substantial interobserver agreement with respect to the overall image quality (κ = 0.69). SSF2 reconstruction improved the image quality of the aortic annulus in the representative case shown in Fig 3.

**Table 6.** Comparison of the image quality scores of scans subjected to standard and SSF2 reconstruction.

|  | R-R<br>interval | Standard | SSF2 | *P* |
| --- | --- | --- | --- | --- |
| Patients with low HR (<60 bpm) | 40% | 2.6 (1.1) | 3.6 (0.7) | <0.001 |
|  | 75% | 3.9 (0.9) | 4.0 (0.7) | 0.540 |
| Patients with intermediate HR (60-69 bpm) | 40% | 2.1 (0.9) | 3.5 (0.6) | <0.001 |
|  | 75% | 2.9 (0.8) | 3.6 (0.7) | 0.003 |
| Patients with high HR (>70 bpm) | 40% | 2.5 (0.7) | 3.7 (0.4) | <0.001 |
|  | 75% | 2.2 (0.7) | 3.2 (0.6) | <0.001 |

HR = heart rate, bpm = beats per minute. Values are the mean (SD).

**Fig 3.**
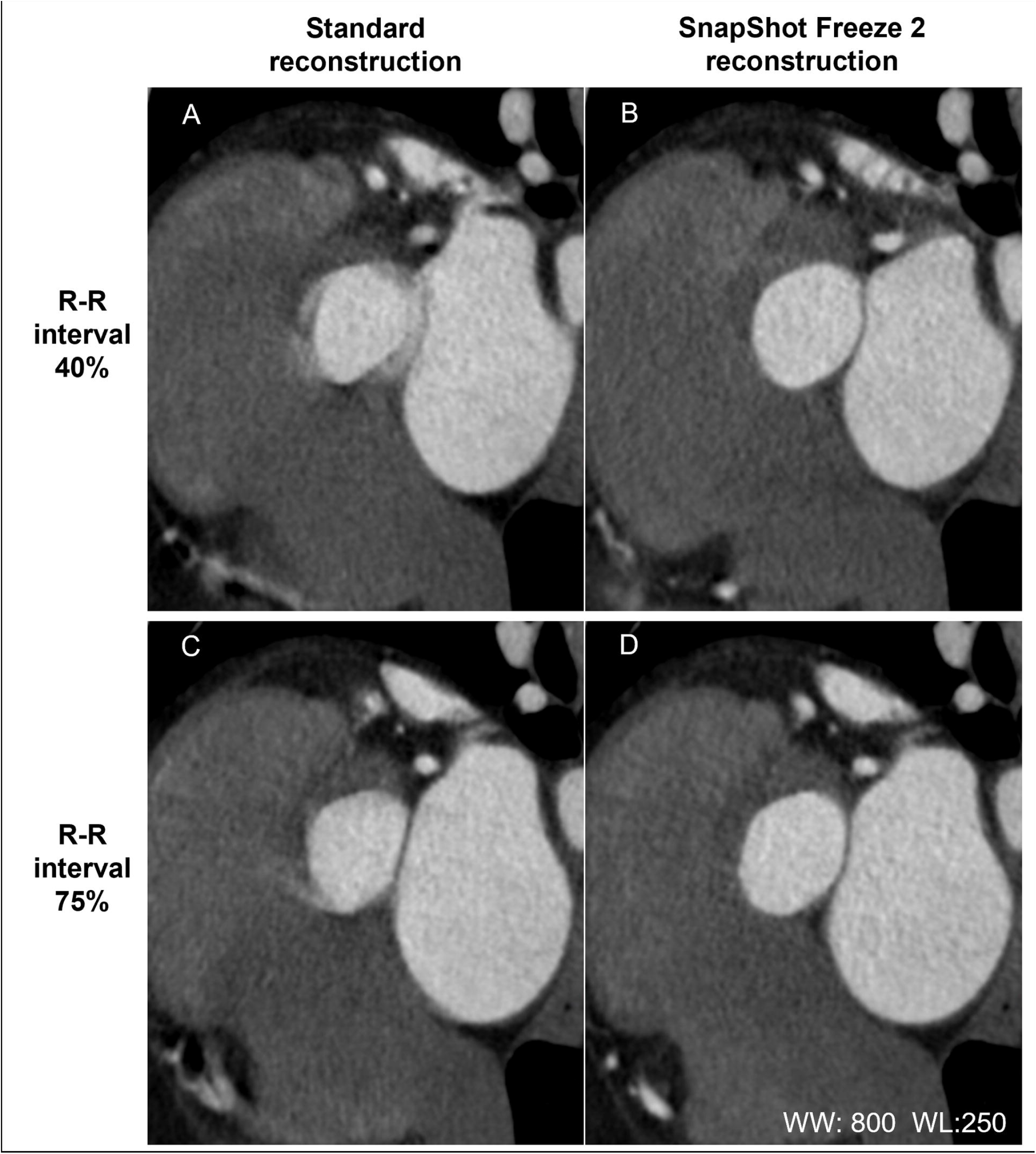
Clinical image of SSF2. In their 80s (heights = 157 cm, body weight = 58 kg, body mass index = 23.5 kg/m^2^, heart rate during the scan = 116 bpm (atrial fibrillation). (A) and (C): MPR images of the aortic annulus (R-R interval = 40% and 75%) using standard reconstruction. The visualization scores for A and C were 1 and 2, respectively. and (D): After SSF2 reconstruction, both visualization scores were 4. The evaluable image quality improved. WW = window width; WL = window level

## Discussion

Our study demonstrates that the second-generation whole-heart motion correction algorithm (SSF2) was superior to standard reconstruction with respect to the image quality of pre-TAVI cardiac CT scans acquired for the evaluation of the aortic annulus.

At R-R 40%, SSF2 reconstructed images received significantly higher image quality scores than did standard reconstruction regardless of the patients’ heart rate (p < 0.001). At R-R 75%, in patients with an intermediate and high heart rate was the visualization score higher for SSF2-than standard reconstructed images. At R-R 40% and R-R 75%, SSF2 strongly tended to yield higher image quality scores than did standard reconstruction. Consequently, SSF2 reconstruction raised the image quality significantly, especially in patients with a high heart rate or a 40% R-R interval.

The earlier vendor-specific motion correction algorithm (SSF1) was designed to address coronary motion artifacts on cardiac scans. It was primarily indicated for coronary imaging and was shown to improve the image quality and diagnostic accuracy of scans performed for the detection of significant coronary stenosis, especially in patients with a high heart rate [12-19]. The SSF2 algorithm extends motion correction to include the whole heart. It is expected to be useful for imaging of not only the coronary arteries but also of other non-coronary intracardiac structures such as the cardiac valves.

Earlier studies that applied SSF2 reconstruction to images of the coronary arteries, of heart-and valve structures, and of the great vessels showed that the image quality was significantly improved by the algorithm and the number of non-evaluable scans was lower than of images subjected to standard-or SSF1 reconstruction [20, 21].

Our study focused on the aortic annulus; it indicates that SSF2 yielded higher motion artifact correction in the whole heart.

Others [29] who applied SSF1 to cardiac CT for aortic annulus measurements reported that it significantly improved the image quality of systolic CT datasets. We examined the effect of SSF2 in a wide range of heart rates and showed that it is useful for the evaluation of the aortic annulus not only in the systolic-but also in the diastolic phase. Our findings suggest that SSF2 reconstruction reduces aortic valve motion artifacts throughout the cardiac phases.

SSF2 reconstruction was not useful at R-R interval 75% in patients with a low or intermediate heart rate. At those heart rates and cardiac phases, the temporal resolution on electrocardiogram-gated scans may be sufficient and motion artifacts may not be inherent. SSF1-and SSF2 reconstruction may be useful in patients with a high heart rate and for scans with low temporal resolution [12, 13, 20, 21]. Our findings suggest that SSF2 is as useful as SSF1 in patients with a high heart rate.

Although cardiac CT is the reference standard for the workup of TAVI candidates scheduled for an investigation of the aortic root [1, 2], motion artifacts reduce the accuracy of aortic annulus sizing and directly impact on patient outcome after TAVI procedure [2, 7-9]. As a result of evaluating the dispersion between the two reconstructions with respect to the sizing of the aortic annulus, SSF2 was significantly smaller than standard regardless of the patients’ heart rate or R-R interval. For TAVI planning, we still tend to use systolic imaging for the measurements [2, 4, 29, 30] and the aortic annulus seems to be better delineated when SSF2 is used. Therefore, SSF2 may contribute to improving the accuracy of sizing of the aortic annulus.

As renal dysfunction is relatively common in elderly patients scheduled for TAVI, a low-contrast protocol is recommended [36]. SSF2 reconstruction may be appropriate in TAVI candidates with renal dysfunction because it not only improves the image quality but also reduces the need for rescanning.

To avoid the potential impact of SSF2 reconstruction on quantitative measurements of the ERD, we measured the CT number in the ascending aorta, the image noise, and the CNR on SSF2 reconstructed images. We found that CNR was equivalent between scans subjected to standard-or SSF2 reconstruction irrespective of the patients’ heart rate, confirming that SSF2 corrected only the motion artifacts and that it did not affect other parameters.

Our study has some limitations. We only focused on the aortic annulus and did not investigate the effect of SSF2 on other cardiac structures such as the coronary arteries. Areas of calcification were excluded from our quantitative evaluation because their CT attenuation fluctuates significantly. Severe aortic valve calcification could reduce the sizing accuracy of the aortic annulus and further study is required to evaluate whether SSF2-is superior to standard reconstruction in patients with severe aortic valve calcification. Lastly, we did not investigate the relationship between SSF2 and the radiation dose. Additional studies are underway to determine whether the robustness of SSF2 reconstruction allows lowering the preset padding range prior to scanning, thereby minimizing the required radiation dose.

## Conclusions

In conclusion, our findings suggest that the SSF2 algorithm was superior to standard reconstruction because it improved the image quality and reduces motion artifacts especially in patients with a high heart rate or a 40% R-R interval. These findings may help SSF2 improve the accuracy of sizing of the aortic annulus prior to TAVI.

## Data Availability

All data produced in the present work are contained in the manuscript.

file:///C:/Users/yoriakimatsumoto/Desktop/SSF2%20aortic%20annulus/6-1Plos%20One/Supporting%20Information.htm

## Abbreviations

bpm: beats per minute
CT: computed tomography
CNR: contrast-to-noise ratio
ERD: edge rise distance
MPR: multiplanar reconstruction
ROI: region of interest
SD: standard deviation
SSF: SnapShot Freeze
TAVI: transcatheter aortic valve implantation

## Supporting information

**S1 Table. Raw data for each group**. (XLSX)

